# Discontinuity of care and trust in usual physician among patients with systemic lupus erythematosus

**DOI:** 10.1101/2024.03.13.24304255

**Authors:** Yu Katayama, Yoshia Miyawaki, Kenta Shidahara, Shoichi Nawachi, Yosuke Asano, Eri Katsuyama, Takayuki Katsuyama, Mariko Takano-Narazaki, Yoshinori Matsumoto, Nao Oguro, Nobuyuki Yajima, Yuichi Ishikawa, Natsuki Sakurai, Chiharu Hidekawa, Ryusuke Yoshimi, Shigeru Ohno, Takanori Ichikawa, Dai Kishida, Yasuhiro Shimojima, Ken-ei Sada, Jun Wada, David H Thom, Noriaki Kurita

**Author notes:** **Corresponding Author**: Yoshia Miyawaki, MD, MPH, PhD, Department of Nephrology, Rheumatology, Endocrinology and Metabolism, Okayama University Graduate School of Medicine, Dentistry and Pharmaceutical Sciences, 2-5-1 Shikata-cho, Kita-ku, Okayama 700- 8558, Japan.

## Abstract

**Importance:** Patient trust plays a central role in the patient-physician relationship; however, the impact of outpatient visits with a covering physician (covered visits) on the level of trust in usual physician among patients with chronic conditions is unknown.

**Objective:** To determine whether the number of outpatient visits with a covering rheumatologist is associated with patient trust in the usual rheumatologist.

**Design:** Cross-sectional study.

**Setting:** This study used data from the TRUMP^2^-SLE project conducted at five academic medical centers in Japan.

**Participants:** The participants were Japanese adults with systemic lupus erythematosus who met the 1997 revised classification criteria of the American College of Rheumatology.

The enrollment period was February 2020 to October 2021.

**Exposure:** Outpatient visits with a covering rheumatologist in the past year.

**Main Outcomes and Measures:** The main outcome was patient trust in their usual rheumatologist, assessed using the 11-item Japanese version of the modified Trust in Physician Scale (range 0–100). A general linear model with cluster robust variance estimation was used to evaluate the association between the number of outpatient visits with a covering rheumatologist and the patient’s trust in their usual rheumatologist.

**Results:** Of the 515 enrolled participants, 421 patients with systemic lupus erythematosus were included in our analyses.

The median age was 47.0 years, and 87.2% were women. Thirty-nine usual rheumatologists participated in this study. Patients were divided into groups according to the number of outpatient visits with a covering rheumatologist in the past year as follows: no visits (59.9%; reference group), one to three visits (24.2%; low-frequency group), and four or more visits (15.9%; high-frequency group). The median Trust in Physician Scale score was 81.8 (interquartile range 72.7–93.2). Both the low-frequency and high-frequency groups exhibited lower trust in their usual rheumatologist (mean difference: -3.03 [95% confidence interval -5.93 to -0.80], -4.17 [95% confidence interval -7.77 to -0.58, respectively]).

**Conclusions and Relevance:** This study revealed that the number of outpatient visits with a covering rheumatologist was associated with lower trust in a patient’s usual rheumatologist. Further research is needed to address the potential adverse effects of physician coverage on trust in patient’s usual rheumatologist.

**Key Points:** 

**Question:** Is the number of outpatient visits with a covering rheumatologist associated with the loss of trust in usual rheumatologist in patients with systemic lupus erythematosus (SLE)?

**Findings:** This multicenter cross-sectional study which included 421 patients with SLE revealed that the number of outpatient visits with a covering rheumatologist in the past year was associated with lower levels of trust in the usual rheumatologist.

**Meaning:** This study alerts us about the need to prepare for the possible adverse effects of unavoidable outpatient coverage.

## Introduction

Discontinuity of care is a critical process with the potential to adversely affect the quality of patient care. For example, discontinuity of care has been associated with more avoidable hospitalizations and a higher number of procedures.^1,2^ While significant attention has been directed toward inpatient handovers^3^ and transitions related to year-end resident changeovers,^4,5^ the impact of temporary outpatient physician coverage remains an understudied clinical question. The necessity for coverage of outpatient care, in the absence of the usual physician is a common and unavoidable occurrence,^6^ due to factors such as continuing medical education, maternity leave, or unexpected absenteeism caused by illness.^7^ Acting as a surrogate during the usual physician’s absence,^8^ the covering physician assumes identical responsibilities to those of the primary physician.^9^

Systemic lupus erythematosus (SLE), characterized by multiple organ damage, requires life-long outpatient visits to mitigate functional disability and optimize quality of life. Maintaining a good physician-patient therapeutic relationship is important because outpatient rheumatologists are required to evaluate patients with SLE through laboratory testing and adjust complex medications based on changes in their disease activity. Trust in the primary rheumatologist is central to this therapeutic relationship,^10^ and among patients with SLE, trust is associated with excellent medication adherence.^11^

When a patient’s visit requires coverage by another rheumatologist, the covering rheumatologist is expected to maintain the same quality of care. However, the course of SLE may not be static, and the covering rheumatologist may be forced to make decisions about prescriptions and testing in the absence of an established relationship with the patient.^12^ When a patient with SLE experiences an unsatisfactory visit with a covering rheumatologist, it may erode their trust in their primary rheumatologist. However, although the association between trust and discontinuity of care has been examined primarily in European and US primary care settings, the results from quantitative studies have been mixed,^13^ ^14^ and few studies have focused on the discontinuity associated with outpatient physician coverage within the same department.

The aim of this study was to examine the association between the number of outpatient rheumatologist coverage by colleagues within the same department and patient trust in the primary rheumatologist among Japanese patients with SLE using data from the Trust Measurement for Physicians and Patients with SLE (TRUMP^2^-SLE) project, a multicenter cross-sectional study.

## Methods

### Study design and subjects

This was a cross-sectional study using baseline data from the TRUMP^2^-SLE study, a multicenter cohort study currently conducted at five academic medical centers (Showa University Hospital, Okayama University Hospital, Shinshu University Hospital, Yokohama City University Hospital, Yokohama City University Medical Center). Patients ≥20 years old who met the revised criteria in 1997 for the classification of SLE by the American College of Rheumatology (ACR) were included.^15^ Patients who were seen by their usual rheumatologist for the first time were excluded. All patients provided informed consent prior to enrollment. The patients were consecutively recruited between February 2020 and October 2021.

### Exposure

Exposure in the present study was defined as the patient-reported number of outpatient visits by a covering rheumatologist (NVCs) in the past year. A covering rheumatologist was defined as one in the same facility other than the usual rheumatologist. Based on frequency, the NVCs were classified into three categories: (1) no NVCs (2) one to three NVC (low-frequency group), and (3) four or more NVCs (high-frequency group).

### Outcome

The outcome of this study was trust in patients’ usual rheumatologist, using the Japanese version of the 11-item modified Trust in Physicians Scale by Thom.^10,16^ Each item is rated on a 5-point Likert scale ranging from 1 (totally disagree) to 5 (totally agree). Then, the four negatively worded items were reverse coded and the total score of all items was converted to a range of 0 to 100. This scale has demonstrated high internal consistency reliability (Cronbach’s alpha coefficient = 0.91) and criterion-related validity.^10^

### Covariates

Based on previous literature and the clinical expertise of rheumatologists, the potential confounders were determined to be as follows: patient age,^17^ patient sex,^17^ physician age,^18^ physician sex,^18^ job title,^19^ duration of time since diagnosis of SLE,^19^ disease activity, organ damage, emotional health, the duration of the relationship with their usual rheumatologist (categorized as <1 year, 1-3 years, and 3 years or more) and the number of visits to their usual rheumatologist in the past year (categorized as one to three times, four to six times, and seven times or more).^10,18^ As there are only full professor and associate professor job titles in Japan that contains the Kanji character ‘kyouju’, meaning professor in English, and health information from an authority is more likely to be trusted, the job titles were divided into two groups: associate professor or higher and lecturer or lower.^19^ Disease activity was determined by the usual physician using the Systemic Lupus Erythematosus Disease Activity Index 2000 (SLEDAI-2K). Organ damage was determined by the Systemic Lupus Erythematosus International Cooperative Clinic/American College of Rheumatology Disability Index (SDI). Emotional health was determined by the LupusPRO domain and was converted to a 0–100-point score.^20^ Higher emotional health scores indicated better emotional functioning and role emotional.

### Statistical analysis

Patient characteristics were summarized separately by NVCs with continuous variables as median and interquartile range (IQR) and categorical variables as frequencies and percentages. A general linear model was fit to examine the association between NVCs and the trust in the usual rheumatologist. All the above-mentioned covariates were entered into the model. In addition, to examine the possibility that the duration of the relationship with their usual physician may modify the association between NVCs and trust in the physician, their product terms were entered into the model. The presence of the interaction was assessed using the Wald test. Next, a logistic regression model was fit with all the above-mentioned covariates as explanatory variables to explore factors associated with high-frequency NVCs. In both of the models, cluster-robust variance estimation was used with each usual rheumatologist as a cluster unit to address the possibility of outcome similarity (i.e., clustering) for the same usual rheumatologist.^21^ Multiple imputation with chained equations was used to address missing data, assuming that the data were missing at random.^22^ All missing values were multiply-imputed 100 times in the imputation process. These estimates were combined using Rubin’s rule.^22^ All analyses were performed using Stata software (version 17.0; Stata Corp, College Station, TX, USA). Statistical significance was set at p<0.05.

### Ethical considerations

This study was conducted in accordance with the Declaration of Helsinki, and the study protocol was approved by the Graduate School of Medical and Dental Sciences of Okayama University and the Okayama University Hospital Ethics Committee (approval number: 2204-020).

## Results

### Patient and physician characteristics

Of the 515 patients enrolled, 94 were excluded owing to missing or inadequate data on outcomes and exposures, and 421 were ultimately included in the primary analysis. The participant and physician characteristics are shown in Table 1. The median age of the patients was 47 years (IQR 36–57), and 367 (87.2%) were female. The median duration of the disease was 12.6 years (IQR 6.9–20.3), the median SLEDAI-2K score was 4 (IQR 1–6), and the median SDI was 0 (IQR 0–1). Regarding NVCs, 252 (59.9%) had no visits, 102 (24.2%) had low frequency (1-3 visits), and 67 (15.9%) had high frequency ( ≥4 visits). Patients were seen by a total of 39 usual rheumatologists. The median age of the rheumatologists was 43 years (IQR 38–48), and nine (23.1%) were female. Seven (17.9%) rheumatologists were associate professors or higher in job titles.

**Table 1.** Patient and physician characteristics.

| Patient characteristics,<br>n=421 | Total | NVC |  |  | Missing, n |
| --- | --- | --- | --- | --- | --- |
|  |  | 0<br>N=252 | 1 to 3<br>N=102 | 4 or more<br>N=67 |  |
| Patient age, median (IQR) | 47.0<br>(36.0-57.0) | 47.0<br>(36.0-59.0) | 42.0<br>(34.8-50.0) | 51.0<br>(41.0-63.0) | 0 |
| Patient sex, female n (%) | 367<br>(87.2%) | 218<br>(86.5%) | 95<br>(93.1%) | 54<br>(80.6%) | 0 |
| SLEDAI-2K score, median (IQR) | 4.0<br>(1.0-6.0) | 4.0<br>(2.0-6.0) | 4.0<br>(2.0-6.0) | 2.0<br>(0.0-6.0) | 0 |
| SDI score, median (IQR) | 0.0<br>(0.0-1.0) | 0.0<br>(0.0-1.0) | 0.0<br>(0.0-1.0) | 1.0<br>(0.0-2.0) | 0 |
| Disease duration, year, median (IQR) | 12.6<br>(6.9-20.3) | 12.6<br>(7.3-18.6) | 9.3<br>(6.3-18.6) | 17.5<br>(9.7-29.2) | 5 |
| Emotional health domain score, median (IQR) | 79.2<br>(54.2-91.7) | 79.2<br>(62.5-91.7) | 79.2<br>(54.2-87.5) | 66.7<br>(33.3-91.7) | 63 |
| Duration of rheumatologist-patient relationship, n (%) |  |  |  |  | 12 |
| < 1 year | 69<br>(16.4) | 36<br>(14.3%) | 21<br>(20.6%) | 12<br>(17.9%) |  |
| 1 to 3 years | 76<br>(18.1%) | 46<br>(18.3%) | 15<br>(14.7%) | 15<br>(22.4%) |  |
| 3 years or more | 264<br>(62.7%) | 163<br>(64.7%) | 62<br>(60.8%) | 39<br>(58.2%) |  |
| Number of visits to usual rheumatologist, n (%) |  |  |  |  | 9 |
| 1 to 3 times | 52<br>(12.4%) | 33<br>(13.1%) | 15<br>(14.7%) | 4<br>(6.0%) |  |
| 4 to 6 times | 210<br>(49.9%) | 132<br>(52.4%) | 44<br>(43.1%) | 34<br>(50.7%) |  |
| 7 times or more | 150<br>(35.6%) | 82<br>(32.5%) | 41<br>(40.2%) | 27<br>(40.3%) |  |

| Rheumatologist characteristics, n=39 | Total |
| --- | --- |
| Rheumatologist age, median (IQR) | 43.0<br>(38.0-48.0) |
| Rheumatologist sex, female, n (%) | 9<br>(23.1) |
| Job title, n (%) |  |
| Lecturer or lower | 32<br>(82.1) |
| Associated professor or higher | 7<br>(17.9) |

IQR; interquartile range NVC; Number of outpatient visits to covering rheumatologist
SLEDAI-2K; Systemic Lupus Erythematosus Disease Activity Index 2000
SDI; Systemic Lupus Erythematosus International Cooperative Clinic/ American College of Rheumatology Disability Index

### Association between NVCs and trust in one’s usual physician

The median Trust in Physician Scale score was 81.8 (IQR 72.7–93.2) and the mean was 81.6 (standard deviation 13.3). The distribution of Trust in Physician Scale Score is shown in Figure 1. The association between NVCs and trust in the patient’s usual physician is shown in Table 2. In the adjusted model, compared to no NVCs, both low and high frequency NVCs were associated with lower Trust in Physician Scale scores, respectively (adjusted mean difference: -3.01 [95% confidence interval (CI) -5.93 to -0.80] and -4.17 [95% CI -7.77 to -0.58], respectively). We failed to demonstrate evidence of a global interaction between the association between NVCs and Trust by duration of the relationship with the usual rheumatologist and NVCs (*P* for interaction = 0.138).

**Figure 1.**
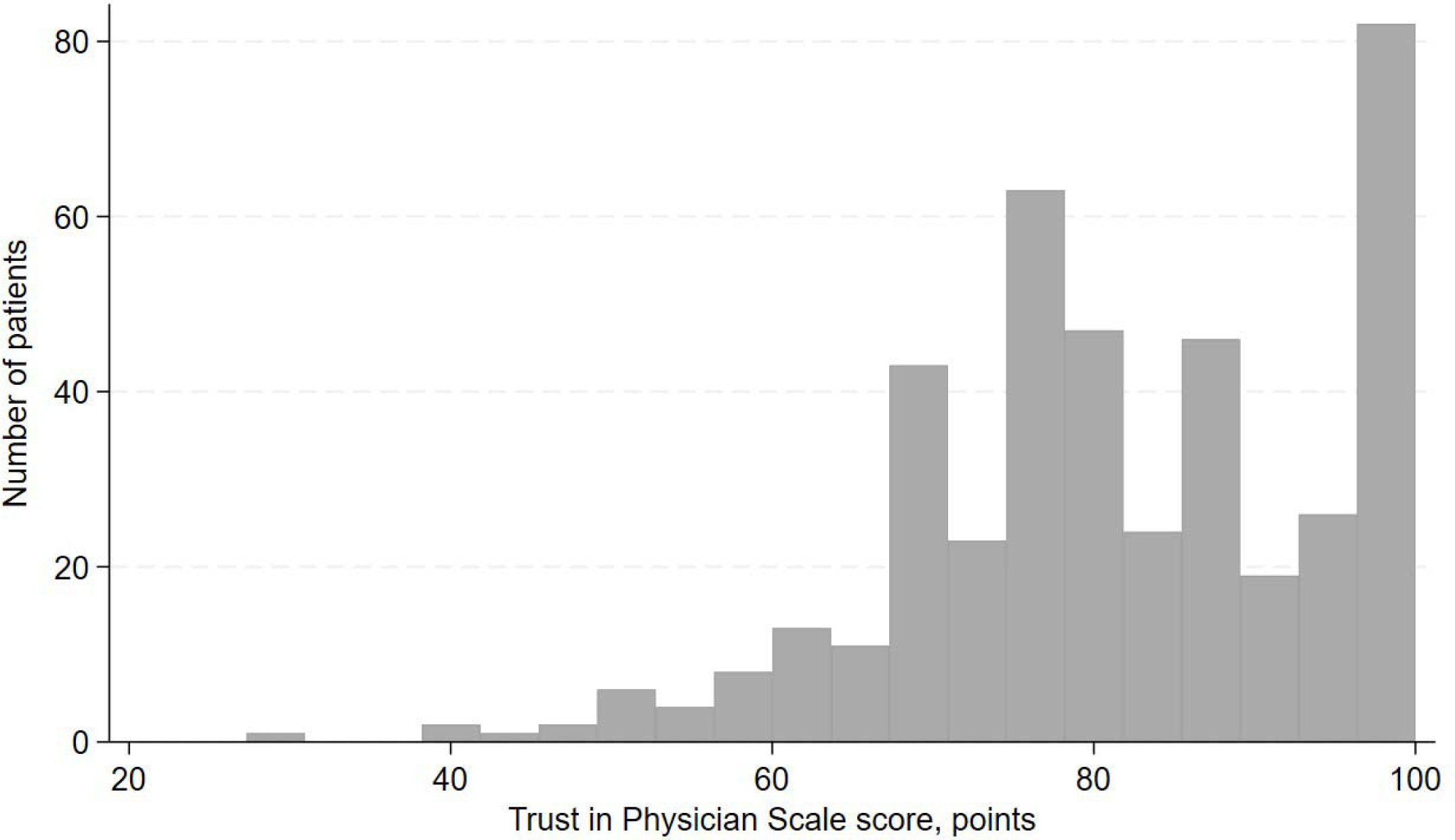
The distribution of Trust in Physician Scale Score The median Trust in Physician Scale score was 81.8 (IQR 72.7–93.2) and the mean was 81.6 (standard deviation 13.3).

**Table 2.** The association between NVCs and trust in one’s usual rheumatologist.

| The number of visits to<br>covering rheumatologist | Crude |  | Adjusted |  |
| --- | --- | --- | --- | --- |
|  | β coefficient<br>[95%CI] | P-value | β coefficient<br>[95%CI] | P-value |
| No visit | Reference |  | Reference |  |
| 1 to 3 visits | -2.53<br>[-5.51 to 0.45] | 0.094 | **-3.01**<br>**[-5.93 to -0.80]** | **0.044** |
| 4 visits or more | **-5.84**<br>**[-9.54 to -2.14]** | **0.003** | **-4.17**<br>**[-7.77 to -0.58]** | **0.024** |

NVC; Number of outpatient visits to a covering rheumatologist
General linear models with cluster-robust variance with usual rheumatologists as the unit were fit to address the clustering of
outcomes by the rheumatologists.
Adjusted models included patient age, sex, rheumatologist age, sex, job title, disease duration, SLEDAI, SDI, emotional health,
duration of rheumatologist-patient relationship, and number of visits to their usual rheumatologist

### Factors associated with high frequent NVCs

The exploratory associations between high-frequency NVCs and covariates are shown in Table 3. Greater organ damage (per 1 point higher SDI: aOR 1.27, 95%CI 1.06–1.52) and more frequent visits to the usual physician (one to three times vs. four to six times: aOR 3.94, 95%CI 1.48– 10.48 / one to three times vs. seven or more times: aOR 3.86, 95%CI 1.46–10.16) was positively associated with a high frequency of NVCs. On the other hand, better emotional health (per point higher: aOR 0.98, 95%CI 0.97–0.99) and higher disease activity (per 1 point higher SLEDAI- 2K: aOR 0.90, 95%CI 0.84–0.97) were inversely associated with a high frequency of NVCs.

**Table 3.**
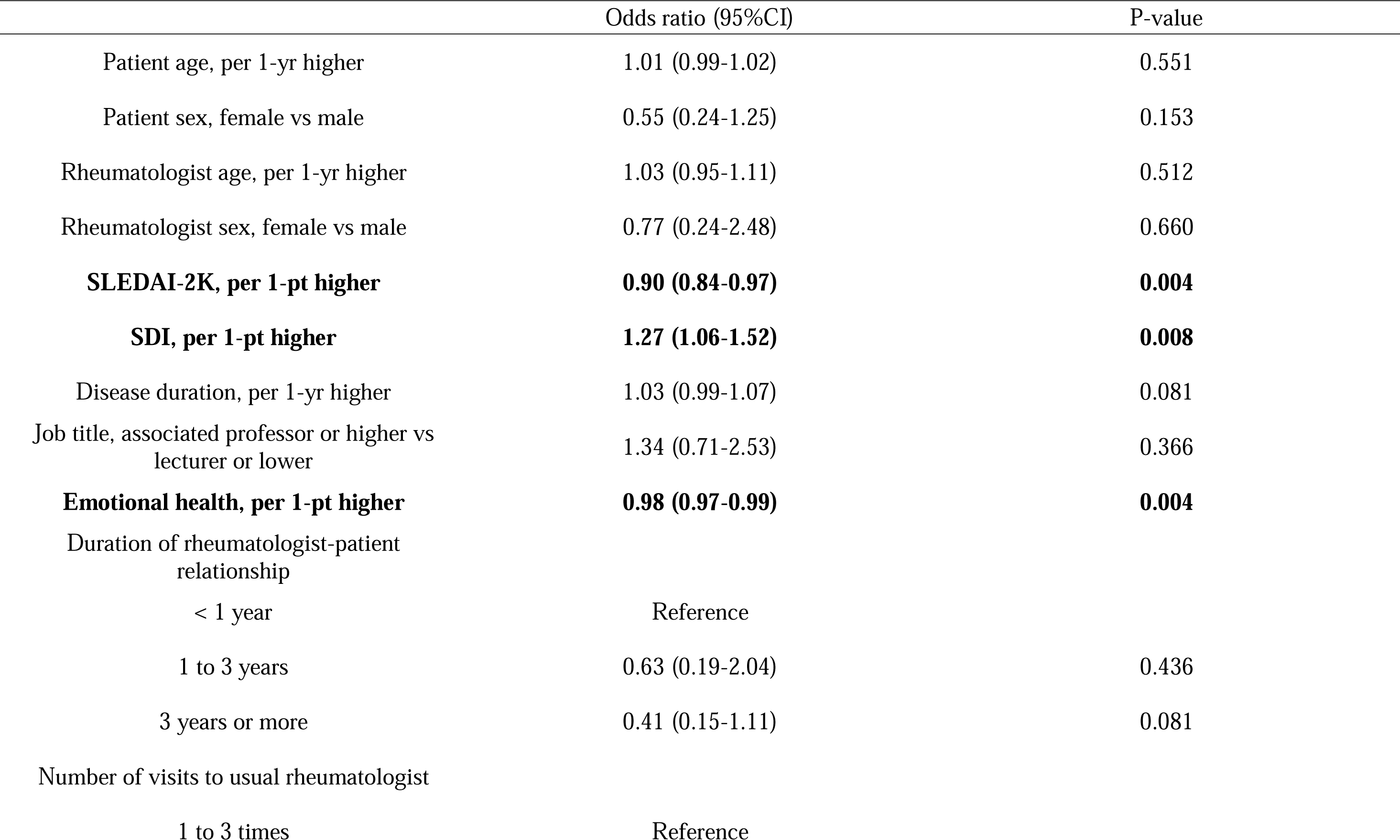
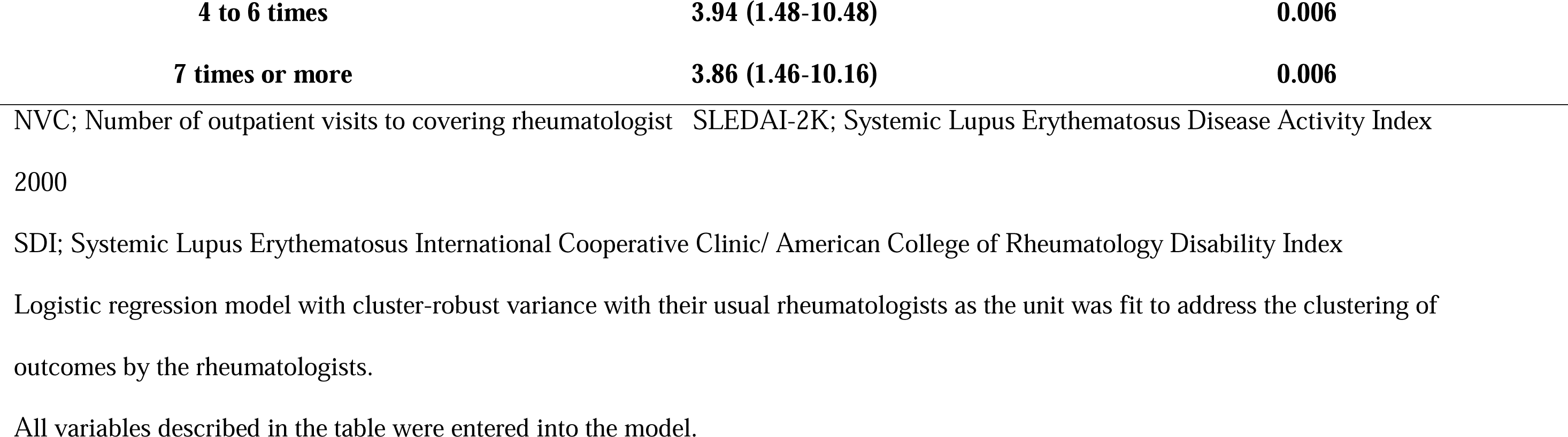
Patient and usual rheumatologist factors associated with high frequent number (≥4 times/year) of outpatient visits covering.

## Discussion

The present study showed that more frequent NVCs with a covering rheumatologist were associated with trust in the usual rheumatologist among patients with SLE. The study found a higher frequency of NVCs was associated with greater organ damage, lower emotional health, and greater number of visits to the usual rheumatologist in the past year.

Few studies have investigated the effects of outpatient visits with a covering physician on the patient relationship with their usual physician among patients with a chronic disease. However, some studies have helped us to understand the findings of our study. First, rheumatologists can understand the possible mechanisms of the loss of trust with increased NVCs by revisiting the level of trust in their physicians. For example, if the usual rheumatologist’s planned care (e.g., performing laboratory tests and explaining their results, or changing the dosage or class of medications) is not communicated to a covering rheumatologist, the patient may feel that the rheumatologists’ words and actions are not credible.^23^ On the other hand, a covering rheumatologist may be hesitant to change medications or order additional testing, preferring to defer to the patient’s usual rheumatologist, which could result in a delayed response to changes in the patient with SLE. In addition, if changes in care are not communicated clearly to the usual rheumatologist, the patient may feel the usual rheumatologist is less informed and therefore less capable of providing the best care. The finding that patients with lower emotional health due to depression and/or anxiety are due to those patients requesting visits more urgently than can be scheduled with their usual rheumatologist.

The clinical implications of this study are noteworthy. First, rheumatologists need to be aware of the magnitude and meanings of frequent NVCs on the patients’ trust in their usual rheumatologists. The magnitude of the 4.17-point lowering of trust scores associated with high frequency (≥4 times/year) NVCs is equivalent to the magnitude (4.30 points) of the impact of past misdiagnosis experiences on trust lowering in the current usual physicians.^10^ Second, rheumatologists need to establish preventive measures against loss of trust resulting from physician coverage. For example, as in the case of computerized tools,^24^ medical facilities would be encouraged to implement a semi-automatic system for handovers of planned laboratory testing, patient explanations, and prescriptions to a covering physician. Alternatively, they could also consider operating an electronic record or a card to be shared whenever any change in patient care occur.^6^ More fundamentally, a system that prevents physician coverage would be ideal. For example, if a rheumatologist takes a day off in advance, a patient scheduled to visit on that day should be notified beforehand to change their appointment date or accept an examination by a covering rheumatologist, thereby avoiding patient surprise or disappointment when they enter an examination room and meets an unfamiliar physician.

This study had several strengths worth mentioning. First, this was a multicenter study; Therefore, the observed effects of rheumatologists’ NVCs on patients’ trust are generalizable to similar academic medical centers, instead of reflecting the specific medical structure of a particular institution’s department. Second, we adjusted for the physician-level clustering of trust with appropriate multilevel analyses by merging data both from usual rheumatologists and from a large number of their patients.^21^ As a result, the observed association between rheumatologists’ NVCs and trust was not reflective of a particular rheumatologist’s behavior.^19^ Third, the racial/ethnic homogeneity means that race/ethnicity is not a complicating factor.^25^

Several limitations should be noticed in this study. First, the possibility of reverse causation cannot be excluded because of the cross-sectional study design. Some patients may have asked to see a covering rheumatologist because they had less trust in their usual rheumatologist. Second, we were unable to survey the reasons for the physician coverage. In addition to the usual rheumatologist’s reasons for arranging coverage, patients may have visited another rheumatologist on a day other than the day of their appointments for patient-related reasons. For example, patients may have rescheduled their appointments on a day when their rheumatologists were not available, or they may have visited another rheumatologist because of unexpected illness. Third, the generalizability of the findings of this study, which were conducted at academic medical centers affiliated with many rheumatologists, may be limited. For example, in the rheumatology departments of city hospitals or private clinics, where only one rheumatologist is often affiliated, rheumatologists must rely on non-rheumatology colleagues or locum tenens for their coverage.^7^ ^26^

In conclusion, frequent outpatient NVCs by a covering rheumatologist were associated with a lower trust in the using rheumatologist among patients with SLE. Given that occasional time off is sometimes required to sustain a medical practice without burnout, this study alerts us about the need to prepare for possible adverse effects of unavoidable outpatient coverage.

## Acknowledgment

### Author Contributions

Dr Katayama and Kurita had full access to all of the data in the study and take responsibility for the integrity of the data and the accuracy of the data analysis.

*Concept and design*: Katayama, Miyawaki, Kurita. *Acquisition, analysis, or interpretation of data*: All authors. *Drafting of the manuscript*: Katayama, Miyawaki, Kurita.

*Critical revision of the manuscript for important intellectual content*: All authors. *Statistical analysis*: Katayama, Miyawaki, Kurita.

*Obtained funding*: Y.Matsumoto, Kurita

*Administrative, technical, or material support*: Katayama, Miyawaki, Yajima, Yoshimi, Shimojima, Sada, Kurita.

*Supervision*: Miyawaki, Yajima, Yoshimi, Shimojima, Sada, Thom, Kurita.

### Funding/Support

This study was supported by JSPS KAKENHI Grant Numbers 19KT0021 (N. K.) and 22K19690 (N. K.).

### Data Sharing Statement

The dataset analyzed in this paper is available from the corresponding author on reasonable request.

### Additional Contributions

The authors are grateful to all collaborators in the TRUMP^2^-SLE project.

